# Protocol for the development of a meta-core outcome set for stillbirth prevention and bereavement care following stillbirth

**DOI:** 10.1101/2022.10.13.22281030

**Authors:** Kushupika Dube, Elizabeth Ayebare, Danya Bakhbakhi, Carol Bedwell, Savitha Chandriah, Nasim Chaudhry, Ides Chilinda, Angela Chimwaza, Unice Goshomi, Rose Laisser, Tina Lavender, Tracey A Mills, Sudhindrashayana Fattepur, Bellington Vwalika, Sabina Wakasiaka, Jamie J Kirkham

## Abstract

**Introduction:** A stillbirth is the death of a baby before or during birth and accounts for about 14 in every 1,000 births globally with the highest rates seen in Sub-Saharan Africa and South Asia. Stillbirth prevention and bereavement care following stillbirth remains a challenge, particularly in Low-Middle Income Countries (LMiC). One approach to improvement is the prioritisation of women/family-centred care. However, there are a large variety of outcomes measured in stillbirth studies and consensus on the outcomes that matter most to women and families is often lacking, which can impact on the ability to make informed decisions about improved care practices. To help mitigate this problem, a core outcome set (COS) has been developed for stillbirth prevention and another COS has recently been finalised for care after stillbirth. Despite the majority of stillbirths occurring in LMiC involvement in these studies is ‘tokenistic’ and therefore the outcomes may not reflect the needs of parents or communities in these settings. The aim is to develop standard sets of outcomes for use in all interventional studies for stillbirth prevention and bereavement care using participants from predominantly Sub-Saharan Africa and South Asia, where the burden of stillbirth is highest.

**Methods/Design:** This study will involve three stages in the development of the COS: (1) a list of outcomes will be identified from multiple sources, specifically existing reviews of outcomes and a targeted qualitative literature review of studies that have interviewed parents who have experienced stillbirth and healthcare professionals working in this field across Sub-Saharan Africa and South Asia. (2) The list of outcomes will first be reviewed by in-country leads and scored by multiple stakeholder groups in a real-time online Delphi survey. (3) The results of the Delphi will be summarised and discussed at a face-to-face or virtual consensus meeting with representation from all stakeholder groups.

**Discussion:** As well as improving the consistency of outcomes for future research in an LMiC setting, these COS will harmonise with the existing COS in this field developed in a high income setting. The final output will be a global ‘meta-COS’, a recommended set of outcomes that can be used in stillbirth research worldwide.

## Introduction

In 2019, it was reported that an estimated 2 million stillbirths occurred annually with most stillbirths occurring in sub-Saharan Africa and south Asia [1]. In order to observe real improvement in public health outcomes in the regions most impacted by poor perinatal outcomes, it is important that LMiC countries become the generators and not the recipients of research data [2].

To increase the relevance and comparability of results from stillbirth studies, a core outcome set (COS) for stillbirth prevention (COSTIL COS) has been developed to improve the usefulness of research to guide clinical practice in this field [3]. In addition, a COS has been agreed (iCHOOSE COS) for care after stillbirth [4] but is not yet published. Despite the robust methodology used to develop these COS and the inclusion of the three main stakeholder groups (clinicians, researchers and parents) identified by the Core Outcome Set-STAndards for Development (COS-STAD) [5] within the consensus process, the involvement of stakeholders, particularly parents from an LMiC settings has been limited. Specifically, for the published COSTIL COS, it was noted that while some UK clinician participants had experience in practice and research within an LMiC setting, no parents from these regions were included in the study development. Given the burden of global stillbirths in LMiC settings, and the limited amount of involvement of stakeholders from those in these regions, it is unclear whether these existing COS are representative enough to be considered a ‘Global’ COS.

### Aims and Objectives

The aim of this study is to contribute to the rapid development of a COS exclusively to an LMiC setting, relevant to all interventions and care options for use in studies for preventing stillbirth and improving bereavement care following stillbirth. In combination with the findings from the COSTIL and iCHOOSE COS, a recommendation will be made on a ‘meta-COS’ which could be seen as the first global COS for stillbirth studies across the world.

The specific objectives are as follows: to utilise the list of outcomes used in the existing COS studies together with outcomes reported in qualitative literature in an LMiC setting relevant to stillbirth; to review and prioritise outcomes from a health care professional, researcher and parent perspective; and to integrate the outcomes important to all stakeholders in order to ratify the COS.

### Scope of the core outcome set(s)

Two COS will be developed, one for stillbirth prevention and one for bereavement care following stillbirth. Both COS will be developed for research and clinical practice and will consider all interventions and care options for stillbirth care within this scope. These COS will predominantly be developed for use in an LMiC setting but the recommendations from COS developed on related topics (COSTIL and iCHOOSE) within a high-income setting will be compared, and suggestions made on priority outcomes that could be used for stillbirth research more globally.

### Steering Committee Membership

The Study Steering Committee (SCC) will comprise membership from a multidisciplinary team within a NIHR Global Health Research Unit (GHRU) on the Prevention and Management of Stillbirths and Neonatal Deaths in Sub-Saharan Africa and South Asia. The team includes health care professionals, community engagement and involvement representatives including bereaved parents (mothers and fathers) and methodologists (inclusive of a COS development expert) representing six Sub-Saharan African countries (Kenya, Malawi, Tanzania, Uganda, Zambia and Zimbabwe), two countries in South Asia (Pakistan and India) and UK experts with substantial collaborative research experience in an LMiC setting.

### Expert panel

An expert panel of two members has been convened to review and guide the Steering Committee at each stage of the project. Members of the panel include Ben Mol and Danya Bakhbakhi, investigators on the COSTIL and iCHOOSE COS respectively.

## Methods/Design

In the development of this protocol, we adhere to the Core Outcome Set-STAndards for Development (COS-STAD) recommendation [5], and follow the COS-STAP statement (Core Outcome Set-STAndardised Protocol Items) [6], for developing a COS protocol. The study is registered in the Core Outcome Measures in Effectiveness Trials (COMET) database [7].

### Step 1: identification of the long list of outcomes

The list of outcomes for use in the real-time Delphi survey will be generated from the following sources:

1. The 56 outcomes entered into the Delphi study from the COSTIL study [3] and the 108 outcomes entered into the Delphi for the iCHOOSE study [4]
2. Outcomes will also be extracted from a narrative synthesis of the qualitative literature using methods described in a similar review for type 2 diabetes [8]. In brief the review will aim to capture the outcome suggestions from the several hundred qualitative interviews already conducted in an LMiC setting with both parents and health care professionals on the topic of stillbirth [9-11].

### Review of a list of outcomes

The list of all outcomes from both sources will be reviewed by the study authors and separated out into outcomes relevant to the prevention of stillbirth, outcomes related to bereavement following stillbirth, and also by outcome domains used in COSTIL and iCHOOSE.

Following the groupings, the outcome lists will be circulated to the eight country leads (CLs) from Sub-Saharan Africa and South Asia participating in the study. Plain language summaries of each outcome and a summary of Delphi scores (if available) obtained for each outcome from the COSTIL and iCHOOSE COS studies will be also presented. Each CL will then have an opportunity to comment on these descriptions and results through think-aloud interviews [12], a technique also used in the iCHOOSE COS to improve questionnaire development and to refine the list of outcomes [4]. The recorded interviews conducted over ‘Zoom’ will be used to examine how CLs interpret outcome descriptions and to review the relevance (scope) of each outcome with respect to a LMiC setting, and with some knowledge on how outcomes were rated in the two previous related COS studies. The audio recorded comments will be reviewed by a member of the SCC and suggested changes in the wording of outcomes or any suggestion of combining similar outcomes and dropping of outcomes that fall out of scope will be made with the justification for these choices documented. The aim is to reduce the long list of outcomes to ensure the number of outcomes to score in the Delphi survey is manageable.

### Step 2: outcome prioritisation

#### Stakeholder involvement

Stakeholder groups representing health care professionals (e.g. obstetricians, nurse midwives), researchers and parents will be invited to participate in the consensus process (real-time Delphi survey and face-to-face consensus meeting). The same participants will contribute to both the COS for stillbirth prevention and bereavement care following stillbirth.

Participants will be invited through the NIHR GHRU network, previous NIHR Group and through CL contacts. For the Delphi survey, the aim is to recruit a minimum of ten health care professional and parent participants (mother and fathers) from each of the six Sub-Saharan African and two South Asian countries involved with the GHRU network and Group. At least ten researchers across the entirety of the network is the target.

#### Real-time Delphi survey

Stakeholders will be invited to score individual outcomes in a real-time Delphi survey, firstly for stillbirth prevention and then for bereavement care following stillbirth. Real-time Delphi surveys have the prospect of improving the speed and efficiency of gathering opinions on outcomes than standard multi-round approaches [13]. Participants will be asked to rate the importance of each outcome on a 9-point Likert scale (1-3 limited importance, 4-5 important but not critical and, 7-9 critical) following the GRADE (Grading of Recommendations, Assessment, Development and Evaluations) guidelines [14]. On scoring, participants will immediately receive anonymised feedback which will consist of the individuals rating and the distribution of scores for each outcome according to a) each stakeholder group and b) overall across all stakeholder groups. Once feedback is received, participants will have the opportunity to re-review outcomes and change their scores if they wish. At the end, participants may also add any additional outcomes they think important but not already included in the list. Any additional outcomes will be reviewed by the SCC to consider whether these outcomes are relevant and not duplicated. Participants will be reminded by email to re-visit and re-rate outcomes before the survey ends, where they will also be able to score, receive feedback and re-score the additional outcomes if necessary. To maximize participation and to ensure specific groups of potential participants are not excluded, where possible, CLs will provide support with providing technologies into the communities in order to fill in the online survey, as well as providing 1:1 support with the questionnaire. The real-time Delphi survey will be managed through Calibrum real-time Delphi software [15].

#### Consensus Criteria

On completion of the Delphi survey, the results will be summarised according to the pre-specified definition of consensus (Table 1).

**Table 1:** Consensus criteria

| Consensus Classification | Description | Definition |
| --- | --- | --- |
| Consensus in | Consensus that the outcome should be included in the final core outcome set | 70% or more participants scoring as 7–9 (critical) AND < 15% participants scoring as 1–3 (limited importance) in all stakeholder groups |
| Consensus out | Consensus that the outcome should <u>not</u> be included in the final core outcome set | 50% or fewer participants scoring 7–9 (critical) in all stakeholder group |
| Equivocal | Uncertainty about the importance of the outcome | All other responses |

#### Step 3: consensus meeting and final core outcome set development

##### Consensus meeting

The proposal is to organise a face-to-face meeting comprising of the SCC and selected participant members representing similar numbers from all stakeholder groups from LMiC countries across the NIHR GHRU Network. The meeting will coincide with a planned gathering of the Network participants although there will be capacity to switch to an online or hybrid meeting if necessary. The results to all outcomes for stillbirth prevention and bereavement care after stillbirth will be presented separately and in accordance to consensus criteria definitions (Table 1) from the Delphi survey. Where outcomes from the Delphi reached ‘consensus in’ or ‘consensus out’, participants will be invited to briefly discuss and provide more information if they disagree with the inclusion/exclusion of the outcome in the COS. Following discussion, participants of the consensus meeting will re-score the outcome again using the GRADE scale. Where outcomes were equivocal during the Delphi survey, they will be discussed and participants of the consensus meeting will be invited to re-score the outcome. Where voting is required, this will be undertaken anonymously with 80% or more stakeholders needing to rate the outcome as critically important (score 7-9) to be considered as candidate outcomes for the COS.

##### Final core outcome set

At the end of the consensus meeting, the consensus meeting panel will review the proposed outcomes to be included in the COS following the discussions and voting. A final reflective discussion will be undertaken to ensure the outcomes included are pragmatic and feasible to measure in an LMiC setting. Furthermore, if appropriate, we will aim to ensure that there is a good balance of outcomes in the core sets which reflect outcomes related to the mother, offspring and the core areas of healthcare indicated by the outcome domains. If a final COS is not agreed on at the end of the consensus meeting, subsequent online meetings will be considered in order to ratify the final COS.

##### Meta-COS for stillbirth

When similar COS projects are conducted in parallel, the recommended outcomes from the different studies are complementary to one another even if the scope or methods maybe slightly different. The overlapping outcomes across the COS development studies have been dubbed a meta-COS. The findings from our COS study will be compared to the COSTIL and iCHOOSE COS and overlapping outcomes will be referred to as a global meta-COS for use in stillbirth research and clinical practice (prevention and bereavement care) in both a high income and LMiC setting.

##### Ethics/Dissemination

In accordance with Good Clinical Practice (GCP) ethics approval has been obtained from the study sponsor, Liverpool School of Tropical Medicine, UK (reference 21-097). In country ethics was also obtained from each of the eight participating countries; India (Bangalore Medical College & Research Institute, reference: BMCR/PS/85/2022-23), Kenya (University of Nairobi, reference: P422/05/2022), Malawi (Kamuzu University of Health Sciences, reference: P.05/22/3632), Pakistan (Health Research Institute National Bioethics Committee, reference: No.4-87/791/22/2254), Tanzania (Catholic University of Health and Allied Sciences Bugando, reference: CREC/554/2022), Uganda (Makerere University, reference: MAKSHSREC-2022-278), Zambia (University of Zambia, reference: 2728-2022), Zimbabwe (Women’s University in Africa, reference: 03/2022).

The development of these COS will be reported according to the COS-STAR (Core Outcome Set-STAndards for Reporting) guidelines [16]. After publication we will register the COS with CROWN (Core Outcomes in Women’s & Newborn Health) [17] and share our findings across the GHRU Network, the LAMRN (Lugina Africa Midwives Research Network) [18] and other relevant international societies and organisations for wider dissemination.

## Discussion

Over 250 COS have been published but less than 10% (22/256) of these have included participants from Africa [19]. Even those COS that have included LMiC participants, the number involved is often disproportionality small compared to their high income country counterparts, which means LMiC participant opinions may become overshadowed when the results are aggregated using some of the existing COS consensus approaches. While some research priorities seem to be applicable to both LMiC and high income countries, differences may occur between these broad regions which is evident in stillbirth where LMiC countries have the highest burden compared to all other regions in the world.

Reported barriers in conducting research in lower income countries include a lack of time and competing priorities, lack of funding and infrastructure for research and ethical and regulatory obstacles [20]. This proposed stillbirth COS development project for a LMiC setting overcomes many of these challenges because of the capacity building networks that already exist through the NIHR GHRU but also the well-established existing links by the researchers in Sub-Saharan Africa via the Lugina Africa Midwives Research Network (LAMRN) [18]. Moreover, the project aims to partly address the involvement of stakeholders from LMiC which is seen as an evidence gap in COS development [21] and the need to consider choices of outcomes for an LMiC setting, which was the number one most important topic in a trials methodology research priority setting exercise for research in this setting [22].

The project builds on the work of the COSTIL and iCHOOSE COS studies but does not aim to replace these important pieces of research. The project uses methods that aims to utilise what work has already been done on this topic already rather than duplicate it. This approach can be seen as a ‘top-up’ research, a more rapid COS development approach to convert an existing International COS into a more Global COS, whilst still adhering to methodological standards [5].

An advanced limitation of our work is that the consensus process will be conducted in the English language. As part of the ethics application process for some countries, project and patient information sheets in local language were required which will be used in this study. While most of our potential participants have a good understanding of the English language, it is acknowledged that in some cases this might be a barrier to participation. In order to mitigate this problem and where necessary, country leads will offer support with both the technology and the understanding of the materials required to participate within the study.

The concept of a meta-COS is not new and has been discussed in other areas such as paediatric asthma [23] and COVID-19 in hospitalised patients [24]. Such meta-COS result from different groups of researchers working on the same COS unknowing to one another until the studies are published. In this study we will collaborate with both the COSTIL and iCHOOSE working groups and share guidance with other COS developers on methods for improving the inclusivity of LMiC participants within their studies.

## Data Availability

This is a study protocol - no data to date have been collected.

## Author’s contribution

JJK conceived the idea for the project. JJK, KD, EA, DB, CB, SC, NC, IC, AC, UG, RL, SF, TL, TAM, BV and SW were all involved in obtaining ethics for this study. All authors provided suggestions and contributed to the design of the study. JJK prepared the initial protocol manuscript. All authors were involved in revising and approving the final manuscript.

## Acknowledgement

The authors would like to acknowledge Allen Nabisere (Uganda) and Sehrish Toufique (Pakistan) for their contribution in adapting the information sheets and consent and translational work for the respective ethics applications, and also Sudhin Thayyil and Ismita Chhetri for facilitating the workflow to date with the team in India.

## Author’s contribution

CL: Country Lead, COMET: Core Outcome Measures in Effectiveness Trials, COS: Core Outcome Set; COS-STAD: Core Outcome Set-STAndards for Development, COS-STAR: Core Outcome Set-STAndards for Reporting, COS-STAP: Core Outcome Set-STAndardised Protocol Items, COSTIL: Core Outcomes in Stillbirth, CROWN: Core Outcomes in Women’s & Newborn Health, GHRU: Global Health Research Unit, GRADE: Grading of Recommendations, Assessment, Development and Evaluations, iCHOOSE: International Collaboration for Harmonising Outcomes fOr Stillbirth research and care, LAMRN: Lugina Africa Midwives Research Network, LMiC: Low Middle income Country, SCC: Study Steering Committee.

